# 90K/*LGALS3BP* Expression is Upregulated in COVID-19 but Does Not Restrict SARS-CoV-2 Infection

**DOI:** 10.1101/2022.07.18.22277255

**Authors:** Laure Bosquillon de Jarcy, Bengisu Akbil, Johanna Leyens, Dylan Postmus, Greta Harnisch, Jenny Jansen, Marie L. Schmidt, Annette Aigner, Fabian Pott, Robert Lorenz Chua, Lilian Krist, Roberta Gentile, Barbara Mühlemann, Terry C. Jones, Daniela Niemeyer, Julia Fricke, Thomas Keil, Tobias Pischon, Jürgen Janke, Christian Conrad, Stefano Iacobelli, Christian Drosten, Victor M. Corman, Markus Ralser, Roland Eils, Florian Kurth, Leif Sander, Christine Goffinet

**Author notes:** **Corresponding Author Contact Information** Christine Goffinet, Institute of Virology, Campus Charité Mitte, Charité - Universitätsmedizin Berlin, Charitéplatz 1, 10117 Berlin, Germany.

## Abstract

Glycoprotein 90K, encoded by the interferon-stimulated gene *LGALS3BP*, displays broad antiviral activity. It reduces HIV-1 infectivity by interfering with Env maturation and virion incorporation, and increases survival of Influenza A virus-infected mice via antiviral innate immune signaling. Here, we analyzed the expression of 90K/*LGALS3BP* in 44 hospitalized COVID-19 patients. 90K protein serum levels were significantly elevated in COVID-19 patients compared to uninfected sex- and age-matched controls. Furthermore, PBMC-associated concentrations of 90K protein were overall reduced by SARS-CoV-2 infection in vivo, suggesting enhanced secretion into the extracellular space. Mining of published PBMC scRNA-seq datasets uncovered monocyte-specific induction of *LGALS3BP* mRNA expression in COVID-19 patients. In functional assays, neither 90K overexpression in susceptible cell lines nor exogenous addition of purified 90K consistently inhibited SARS-CoV-2 infection. Our data suggests that 90K/*LGALS3BP* contributes to the global type I IFN response during SARS-CoV-2 infection in vivo without displaying detectable antiviral properties.

## Background

SARS-CoV-2 infects host cells via interaction of its spike protein with the ACE2 receptor [1,2]. Therefore, the infectivity of SARS-CoV-2 particles is largely determined by the characteristics of their spike protein. Antiviral strategies targeting its biosynthesis, maturation, and fusion activity may be clinically beneficial. Membrane fusion-mediating viral proteins are targeted by antiviral proteins expressed from interferon (IFN)-stimulated genes (ISGs). 90K (gene name *LGALS3BP*) is a ubiquitously expressed cellular secreted glycoprotein with multiple antiviral activities. Expression of *LGALS3BP* is stimulated by IFNs, resulting in upregulated 90K serum concentrations in individuals with viral infections, including HIV-1 [3– 5], HCV [6,7], hantaviruses [8], and dengue virus [9]. In the context of HIV-1 infection, 90K expressed in virus-producing cells inhibits proper proteolytic processing of the envelope protein precursor and virion incorporation of glycoproteins, resulting in reduction of particle infectivity [10]. 90K has also been suggested to inhibit virion production through inhibition of HIV-1 Gag trafficking [11]. In addition, 90K activates signaling to mount a cell-intrinsic antiviral profile that was essential for survival of experimental influenza virus infection in mice [12]. Furthermore, secreted 90K may promote NK cell activity, CD8^+^ T-cell-mediated cytotoxicity, and cytokine production [13]. Defective IFN signaling, due to inborn mutations in type I IFN-mediated immunity [14] and presence of autoantibodies against type I IFN [15] have been reported as risk factors for critical COVID-19. Given the IFN-stimulated manner of *LGALS3BP* expression and its reported association with viral infections in vivo, including its potential suitability as a prognostic marker for disease progression in HIV-1/AIDS [16], we investigated the expression profile of 90K/*LGALS3BP* in specimens of COVID-19 patients and uninfected individuals and probed for a potential direct antiviral role of 90K against SARS-CoV-2 infection.

## Methods

### COVID-19 Cohort

Hospitalized COVID-19 patients’ blood samples and clinical data were collected at Charité – Universitätsmedizin Berlin in the context of the *PaCOVID-19 Study* [17]. Patients were recruited between March and November 2020. All patients provided a positive SARS-CoV-2 by RT-PCR from respiratory specimens. The study was approved by the ethics committee of Charité (EA2/066/20). Written informed consent was obtained from all patients or legal representatives. In this work, we analyzed 44 COVID-19 patients. 42 provided serum samples and 13 blood samples for peripheral blood mononuclear cell (PBMC) isolation. Most patients were sampled longitudinally (Suppl. Table 1). To minimize confounding issues, patients with conditions known to enhance 90K serum concentrations were excluded from our study: asthma bronchiale [18], history of malignoma within the last ten years [19], Hepatitis B, C, or liver cirrhosis [20], or HIV-1 [10]. Furthermore, we excluded patients with acute herpes zoster as modulation of ISGs is reported in this context [21].

### Cells

Calu-3 (ATCC HTB-55), Caco-2 (ATCC HTB-37), Vero E6 (ATCC CRL-1586), and HEK293T (ATCC CRL-3216) cells were cultured in DMEM (Sigma Aldrich) with 4.5 g/L glucose, supplemented with 10% fetal bovine serum, 1% penicillin/streptomycin, and 1% L-glutamine at 37□°C and 5% CO2. HEK293T/ACE2 cells were additionally supplemented with 13 µg/ml Blasticidin. Caco-2/90K cells were additionally supplemented with 10 µg/ml Puromycin.

PBMCs from COVID-19 patients and healthy controls from anonymized blood donors were isolated from 2-3 ml EDTA whole blood. Samples were mixed 1:1 with PBS and centrifuged on Pancoll (Pan Biotech) for 30 min at 200 x *g*. PBMCs were then washed with PBS. Remaining erythrocytes were lysed with ACK-Lysis Buffer (8,29g NH_4_Cl, 1g KHCO_3_, 0,0367g EDTA, 600ml H_2_O), followed by PBS washing. PBMC pellets were frozen at -20°C.

### Virus, Lenti- and Retroviral Particles

SARS-CoV-2 strain Munich 984 (strain: SARS-CoV-2/human/DEU/BavPat2-ChVir984, NCBI GenBank Acc. No. MT270112.1) was propagated on Vero E6 cells and concentrated using Vivaspin® 20 concentrators (Sartorius Stedim Biotech). Virus stocks were diluted in OptiPro serum-free medium supplemented with 0.5% gelatine and PBS and stored at -80°C. Infectious titer was defined by plaque titration assay.

VSV-G-pseudotyped lentiviral vector particles encoding human 90K were generated by calcium phosphate-based transfection of HEK293T cells with the packaging plasmid pCMV ΔR8.91 [22], the lentiviral transfer plasmid pWPI-puro-90K-myc or pWPI puro (gift of Thomas Pietschmann) and pCMV-VSV-G [23]. The plasmid pWPI-puro-90K-myc was generated by standard molecular cloning. VSV-G-pseudotyped retroviral vector particles encoding human ACE2 were generated by calcium phosphate-based transfection of HEK293T cells with the packaging plasmid MLV gag-pol [24], the MLV-based transfer plasmid pCX4bsrACE2 [25] and pCMV-VSV-G. Vector-containing supernatant was collected 40 and 64 hours post-transfection and subjected to ultracentrifugation through a 20% sucrose cushion. Aliquots were stored at -80°C.

### Infection with Authentic SARS-CoV-2

Calu-3, Caco-2, and HEK293T/ACE2 cells were seeded in 6-well plates at densities of 6×10^5^, 3×10^5^, and 4.5×10^5^ cells/ml, respectively. Cells were infected with SARS-CoV-2 (MOI 0.01). Virus inoculum was removed after 1h, cells were washed with PBS and resupplied with fresh medium. Where indicated, cells were pretreated with Remdesivir, and treatment was continued until the end of the experiment.

### Data Presentation and Statistical Analysis

Bar graphs indicate mean values and error bars indicate standard deviation. Whiskers in box plots indicate minimum to maximum values. Trends over time are displayed using locally weighted scatterplot smoothing (LOESS) estimates.

To determine statistical significance between two independent groups, we used unpaired t-tests and paired t-tests when comparing cases and matched controls, assuming normal distribution. To analyze data with both independent and dependent observations due to longitudinal measurements of an individual, we used mixed effects models.

Based on scRNA-seq data we used these models to compare expression between cases and controls, just as between mild cases, severe cases, and controls regarding ten cell types from PBMCs and 20 cell types from respiratory samples. As we consider all analyses to be exploratory, we did not adjust for multiple testing.

When analyzing independent observations, graphs and statistical analyses were generated using *Graph Pad Prism 9*.*1*.*2*. Further statistical analyses were performed using R (Ripley 2001; Pinheiro et al. 2021; Wickham et al. 2019). The code utilized for the analysis will be available at https://github.com/GoffinetLab/SARS-CoV-2_90K_patient_study.

## Results

### Serum 90K Concentrations are Elevated in COVID-19 Patients as Compared to Healthy Controls

We quantified 90K concentrations in 119 serum samples from 42 hospitalized COVID-19 patients and 42 age- and sex-matched healthy controls (NAKO). Both mean (Figure 1A) and peak (Figure 1B) 90K serum concentrations were significantly higher in patients with SARS-CoV-2 compared to controls (all p<0.0001). Highest 90K concentrations were detected in patients classified WHO 5 (WHO 3 vs. 5, p=0.052 for mean 90K, p=0.007 for peak 90K). Analyzing all samples with respect to the sampling time point, we saw gradually decreasing 90K levels over time with highest levels at early infection stages (linear mixed effects model, p<0.001) and similar rate of decline in patients with mild and severe disease (Figure 1C). Overall, patients with severe COVID-19 (WHO 5-7) had higher 90K serum concentrations than patients with mild COVID-19 (WHO 3-4) when considering symptom onset (linear mixed effects model p=0.008).

**Figure 1.**
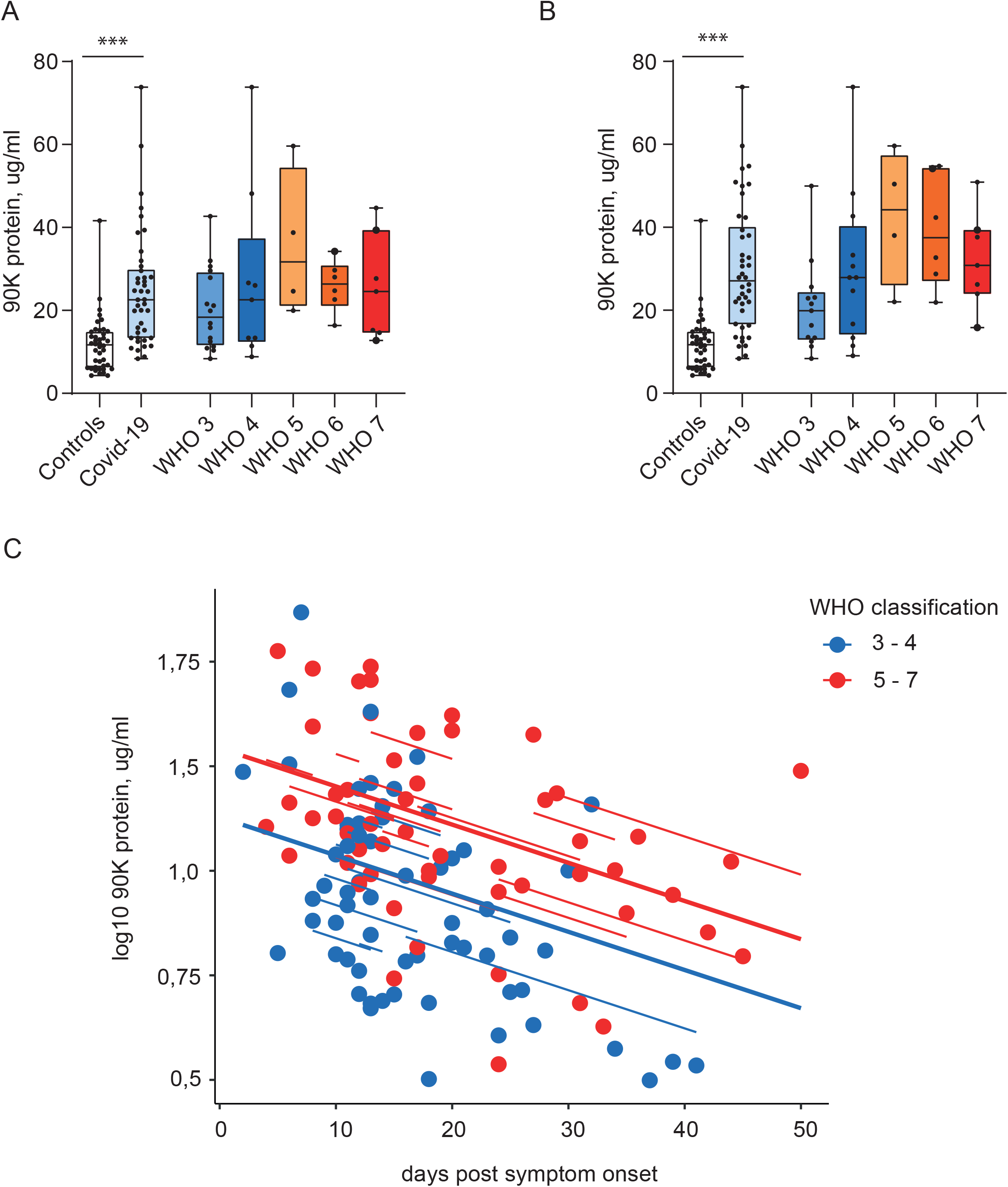
Serum 90K Levels are Elevated in COVID-19 Patients. (A) 90K serum levels in COVID-19 compared to controls. Controls n=42, COVID-19: n=42, WHO 3: n=13, WHO 4: n=12, WHO 5: n=4, WHO 6: n=6, WHO 7: n=7. Multiple ELISA quantifications per patient are presented with the mean, large points indicate deceased patients. Cases vs. controls p<0.0001 and differences between WHO groups (reference WHO 3) were assessed using linear mixed effects models. (B) Peak 90K serum concentrations in COVID-19. Cohort and dot legend identical to Figure 1A. Paired t-test cases vs. matched controls (p<0.0001), unpaired t-test WHO 3 vs. 4 p=0.14, 3 vs. 5 p=0.007, 3 vs. 6 p=0.006, 3 vs. 7 p=0.045. (C) 90K serum concentrations over time in COVID-19 patients, log_10_. n=41/116 (individuals/time points), WHO 3-4: n=24/62, WHO 5-7: n =17/54. 1 our of 42 patients is not depicted for asymptomatic disease course. Log-normalized concentrations were modeled using a linear mixed effects model (marginal R^2^=0.21; conditional R^2^=0.56). The effects of “days post symptom onset” and “WHO classification” were statistically significant (p=0.0004 and p=0.008 respectively). Regression lines are shown, with solid lines indicating the predictions on the population level and dashed lines connecting subjects.

To identify a potential influence of dexamethasone treatment on 90K serum levels, we compared patients treated before and after introduction of dexamethasone as standard of care for COVID-19 patients with oxygen supply [26]. No significant differences between both groups were detectable (Suppl. Figure 1).

Furthermore, we analyzed possible interrelations between 90K serum concentrations, viral RNA concentrations from nasopharyngeal swabs and anti-SARS-CoV-2 IgG/IgA titers and found no association (Suppl. Figure 2 A,B). For demographic aspects, no sex-dependent differences in 90K serum levels were noted (Suppl. Figure 2C), but we recorded higher concentrations in older healthy individuals > 55 y compared to younger individuals (linear mixed effects model p=0.027, Suppl. Figure 2D). This difference was not observed in COVID-19 patients, which displayed enhanced 90K serum concentrations irrespective of age.

### PBMC-associated 90K Protein Concentrations are Reduced in the Context of SARS-CoV-2 Infection

We hypothesized that 90K serum protein originates from peripheral blood cells. Since antiviral functions have been described for intra- and extracellular 90K protein, we aimed at determining if SARS-CoV-2 infection in vivo increases 90K synthesis, or whether 90K secretion or stability may be modulated. Therefore, we quantified cell-associated 90K protein from lysed PBMCs (17 samples from 12 COVID-19 patients) and found reduced concentrations in COVID-19 compared to healthy controls, regardless of disease severity (linear mixed effects model, p=0.014, Figure 2A). This suggests an enhanced secretion or extracellular stability of 90K in COVID-19 rather than an overall enhanced biosynthesis. Furthermore, in contrast to serum 90K, we obtained no evidence for time-dependent changes of cell-associated 90K (Figure 2B) or association with respective serum concentrations in single individuals (Figure 2C).

**Figure 2.**
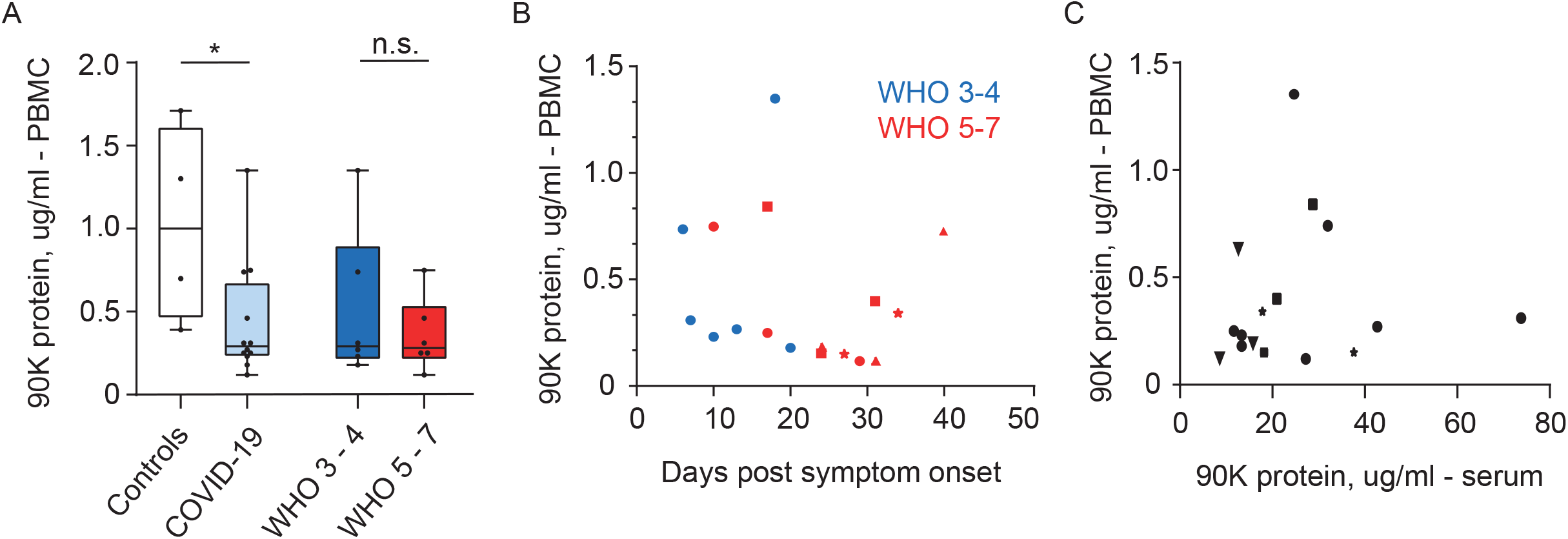
PBMC-associated 90K Protein Concentrations are Reduced in COVID-19. (A) 90K protein in PBMCs, one mean value per individual. Controls: n=4, COVID-19: n=12/17 (individuals/timepoints), WHO 3-4: n=6/6, WHO 5-7: n=6/11. Linear mixed effects model (Controls vs. COVID-19) p=0.01 and (WHO 3-4 vs. WHO 5-7) p=0.39. (B) 90K protein in PBMCs over time in COVID-19. Cohort identical to 2A. Dots show individuals with a single sampling time point. Other symbols show values belonging to longitudinally sampled individuals. (C) 90K protein in PBMCs and sera at identical timepoints in COVID-19 (both samples taken with maximum interval of 48h) n=11/16 (individuals/time points). Dot legend s. 2B.

### *LGALS3BP* mRNA Expression in PBMCs is Unaltered in COVID-19

PBMCs of our COVID-19 cohort (19 samples from n=14 individuals) showed no upregulated *LGALS3BP* mRNA expression compared to healthy controls (Figure 3A) and no time-dependent expression changes (Figure 3B), as judged by qRT-PCR analysis in bulk PBMCs.

In summary, in our cohort of SARS-CoV-2-infected individuals, 90K protein concentrations were enhanced in sera and reduced in PBMCs, while *LGALS3BP* mRNA expression in PBMCs from COVID-19 patients remained unaltered as compared to healthy controls.

**Figure 3.**
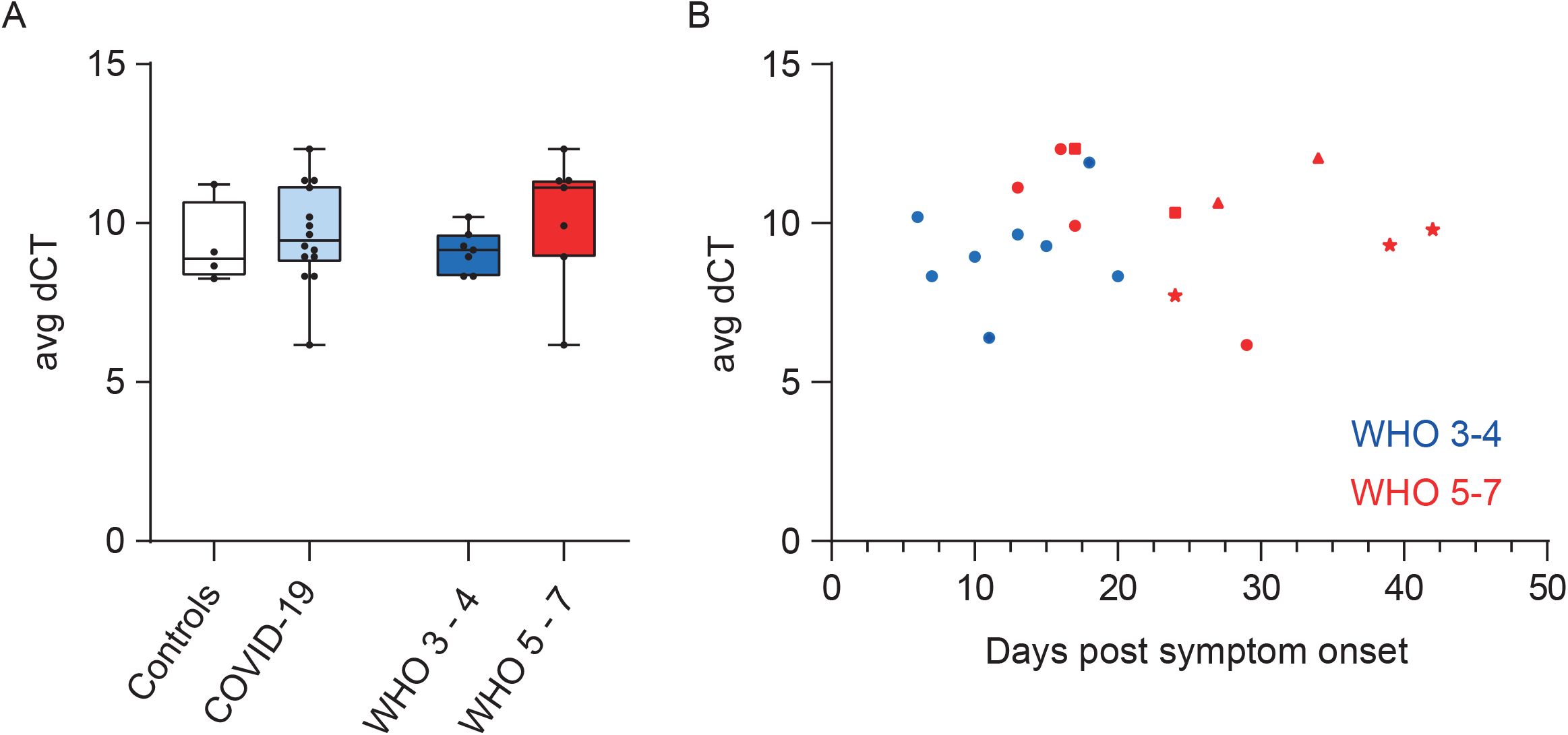
*LGALS3BP* mRNA Expression in PBMCs is Unaltered in COVID-19. (A) *LGALS3BP* mRNA Expression in PBMCs. Controls: n=4, COVID-19 cohort: n=14, WHO 3-4: n=7 and WHO 5-7: n=7, one mean value per individual. (B) *LGALS3BP* mRNA expression in PBMCs over time. Controls: n=4, COVID-19 cohort: n=14/19 (individuals/time points), WHO 3-4: n=7/8, WHO 5-7: n=7/11.

### Single Cell RNA-sequencing Data Reveal Upregulated *LGALS3BP* mRNA Expression Specifically in Monocytes, Dendritic Cells and Plasmablasts of COVID-19 Patients

To identify potential cell type-specific *LGALS3BP* mRNA expression patterns in individual cell types of PBMCs of COVID-19 patients that may be undetectable in our whole-PBMC analysis (Figure 3), we analyzed published scRNA-seq datasets from a Dual Center Cohort Study [27]. As the scRNA-seq data of both cohorts had been generated by different experimental approaches, we analyzed each cohort separately.

In both COVID-19 cohorts, we detected an overall increase of *LGALS3BP* expression in several monocyte fractions compared to healthy controls (Figure 4, Suppl. Figure 7). In cohort B, *LGALS3BP* expression levels were, in addition, significantly elevated in dendritic cells and plasmablasts (mixed linear regression models, p<0.001). Overall, expression was highest at early infection stages and decreased after symptom onset (p<0.01, Figure 4, Suppl. Figure 7). Interestingly, we identified low to absent *LGALS3BP* expression levels in CD163^hi^ and HLA-DR^lo^ S100A^hi^ monocytes of cohort A in severe COVID-19 (Figure 4A) while patients with mild COVID-19 showed elevated levels compared to controls. However, this finding was not reproduced in cohort B.

**Figure 4.**
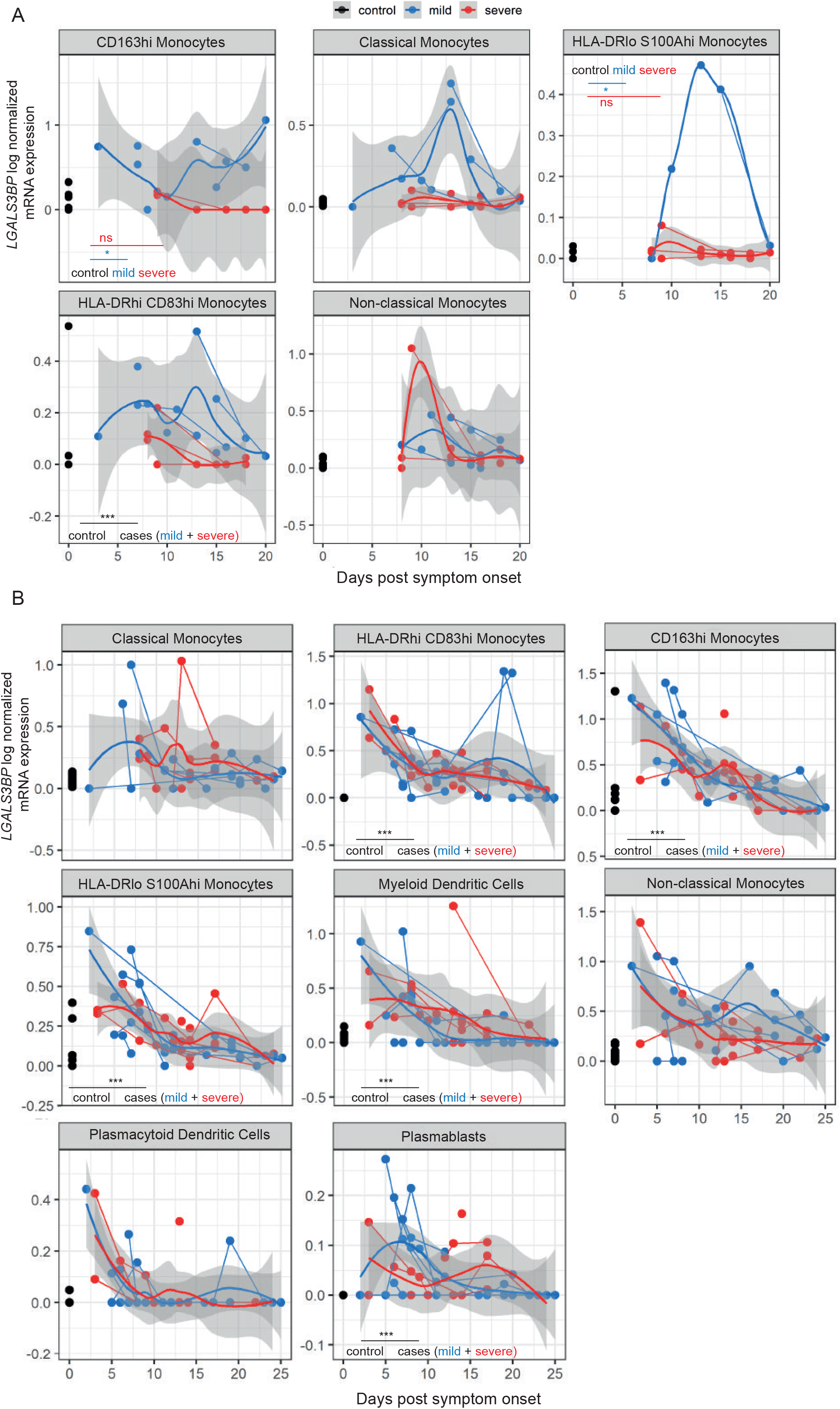
*LGALS3BP* mRNA Expression is Upregulated Specifically in Monocytes, Dendritic Cells and Plasmablasts of COVID-19 Patients. **A**: Cohort A, **B**: Cohort B *LGALS3BP* log normalized mRNA expression over time. Uninfected controls (black), “mild” COVID-19 (blue) and “severe” COVID-19 (red). Thick lines indicate smoothed population trends based on LOESS estimate, thin lines connect subjects. Shaded areas indicate 95% confidence interval (CI) for the LOESS estimate. (A) CD163^hi^ monocytes control: n=19/19, mild: n=7/10, severe: n=4/6 (individuals/time points), classical monocytes control: n=22/22, mild: n=6/13, severe: n=10/14, HLA-DR^Io^ S100A^hi^ monocytes control: n=8/8, mild: n=4/5, severe: n=10/14, HLA-DR^hi^ CD83^hi^ monocytes control: n=9/9, mild: n=6/13, severe: n=10/12, non-classical monocytes control: n=22/22, mild: n=6/10, severe: n=6/13. (B) Classical monocytes - control: n=13/13, mild: n=7/17, severe: n=8/20, HLA-DR^hi^ CD83^hi^ monocytes - control: n=12/12, mild: n=8/22, severe: n=9/28, CD163^hi^ monocytes - control: n=13/13, mild: n=8/21, severe: n=9/26, HLA-DR^Io^ S100A ^hi^ monocytes control: n=13/13, mild: n=8/22, severe: n=9/28, mDCs control: n=13, mild: n=8/22, severe: n=9/23, non-classical monocytes - control: n=13/13, mild: n=8/22, severe: n=9/23, plasmacytoid dendritic cells (pDCs) control: n=12/12, mild: n=8/21, severe: n=9/24, plasmablasts control: n=12, mild: n= 8/22, severe: n=9/28.

### Expression of *LGALS3BP* mRNA is Not Upregulated in Respiratory Samples from COVID-19 Patients

We analyzed the expression of *LGALS3BP* in scRNA-seq data from 32 respiratory samples originating from a previous scRNA-seq study [28]. In contrast to our findings in PBMCs, *LGALS3BP* expression was not increased, neither in epithelial nor in immune cell types of respiratory COVID-19 samples (Suppl. Figure 4).

### SARS-CoV-2 Infection Fails to Induce *LGALS3BP* Expression in Lung- or Colon-derived Cell Lines

To elucidate SARS-CoV-2’s ability to induce *LGALS3BP* expression, we infected Calu-3 and Caco-2 cells with authentic SARS-CoV-2. qRT-PCR revealed a transient, 3-fold and absent induction of *LGALS3BP* mRNA expression, respectively (Figure 5). In line with SARS-CoV-2’s ability to prevent efficient recognition by cell-intrinsic innate immunity [29], no upregulation of *IFIT1* mRNA, a prototypic ISG, was detectable. In contrast, treatment with IFN-α2a induced expression of *LGALS3BP* in both cell lines as expected, although to lower magnitudes than *IFIT1* (Figure 5).

**Figure 5.**
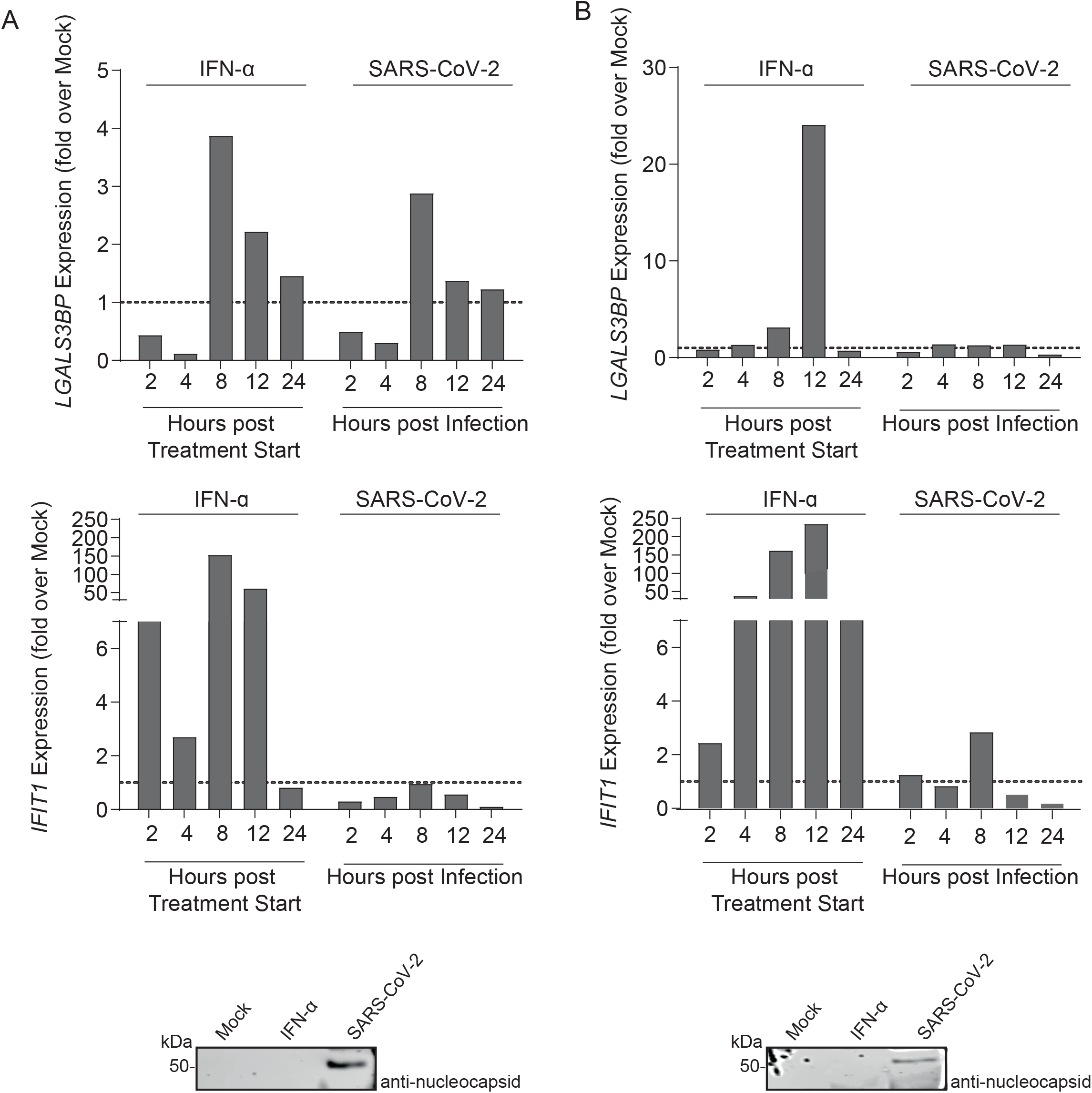
SARS-CoV-2 Infection Fails to Induce *LGALS3BP* Expression in Lung- or Colon-derived Cell Lines. Calu-3 (A) and Caco-2 cells (B) were treated with 500 IU/ml IFN-α2a or infected with SARS-CoV-2 (MOI 0.01). Cells were harvested 2-24h after IFN-α2a treatment or SARS-CoV-2 infection for *LGALS3BP* and *IFIT1* mRNA qRT-PCR. Values are normalized to mock-infected, untreated cells for each timepoint. 24h post infection or IFN-α2a treatment, cells were harvested for immunoblotting.

### Heterologous 90K Expression Mildly Inhibits SARS-CoV-2 Infection in ACE2-expressing HEK293T Cells but Not in Caco-2 Cells

To identify a potential antiviral activity of 90K, we conducted experiments in HEK293T cells that are devoid of detectable 90K expression [10,30] and therefore suitable for heterologous overexpression of 90K. SARS-CoV-2 RNA concentrations in the culture supernatant of infected ACE2-expressing HEK293T cells were reduced 2.5-fold by expression of 90K-myc. Intracellular spike and nucleocapsid expression was diminished 1.7-fold and 3.3-fold, respectively (Figure 6A). Efficient 90K expression was confirmed by immunoblotting.

**Figure 6.**
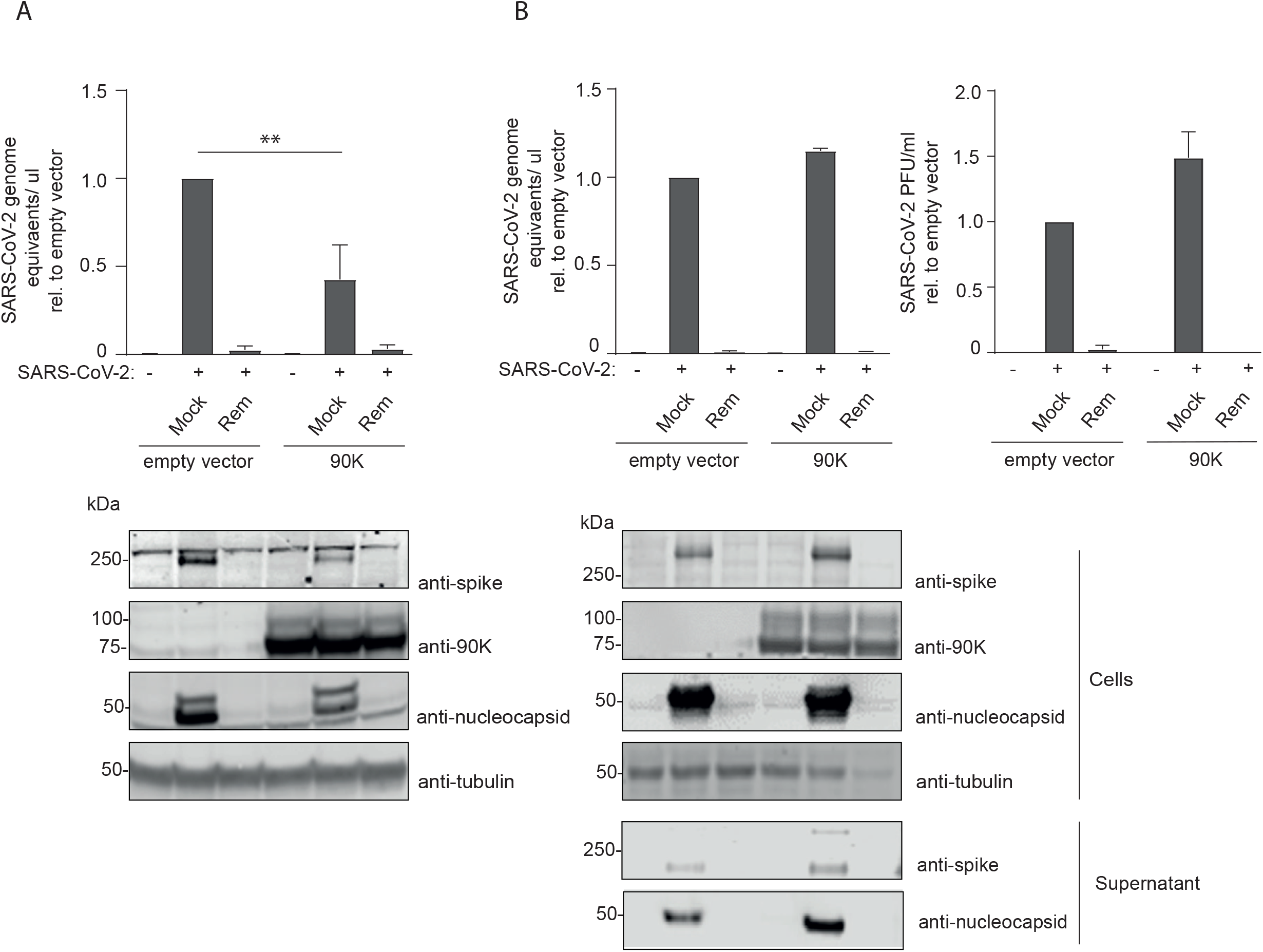
Heterologous 90K Expression Mildly Inhibits SARS-CoV-2 Infection in ACE2-expressing HEK293T Cells but Not in Caco-2 Cells. (A) SARS-CoV-2 genome equivalents from qRT-PCR after SARS-CoV-2 infection of HEK293T/ACE2 cells. Cells were mock-infected or infected in the absence or presence of Remdesivir. 90K-myc was expressed via transient transfection. 24 hours post SARS-CoV-2 infection, supernatants were harvested for quantification of SARS-CoV-2 genome equivalents. Statistical significance was determined using an unpaired t-test (p=0.007). Immunoblot of HEK293T/ACE2 cell lysates using indicated antibodies. (B) SARS-CoV-2 genome equivalents from qRT-PCR and infectivity from plaque assays upon SARS-CoV-2 infection of Caco-2 cells. 90K-myc expression in Caco-2 cells was achieved by lentiviral transduction. 24 hours post SARS-CoV-2 infection, supernatant was harvested for quantification of SARS-CoV-2 genome equivalents and released infectivity. Results are derived from two to three independent experiments. Immunoblot of Caco-2 cell lysates and supernatants using indicated antibodies.

To verify these findings in a SARS-CoV-2-susceptible cell line with endogenous ACE2 expression and undetectable endogenous 90K levels by immunoblotting, we stably transduced Caco-2 cells with empty vector or 90K-myc. In contrast to our finding in HEK293T/ACE2 cells, overexpression of 90K in Caco-2 cells neither reduced the concentration of viral RNA, nor diminished the expression of nucleocapsid and spike in SARS-CoV-2 producing cells (Figure 6B) while pre-treatment with the viral polymerase inhibitor Remdesivir efficiently reduced viral RNA in the supernatant as well as the synthesis of nucleocapsid, as expected.

To analyze the effect of exogenously added 90K protein on SARS-CoV-2 infection, we pretreated HEK293T/ACE2 and Caco-2 cells with 90K or the D2 domain of 90K given its ability to promote cell adhesion (Capone et al. 2021). Pretreatment of cells, or treatment of virus stocks with purified 90K protein or D2 domain of 90K failed to modulate release and infectivity of SARS-CoV-2 particles (Suppl. Figure 6).

## Discussion

Our findings of upregulated 90K serum levels in COVID-19 with highest levels in severe disease courses are in line with proteomic and serological findings from hospitalized COVID-19 patients [31,32]. Overall reduced PBMC-associated 90K protein levels in SARS-CoV-2 infected individuals and, conversely, upregulated *LGALS3BP* mRNA in monocytes, plasmablasts, and dendritic cells hint towards a complex, compartment- and cell type-specific regulation of 90K expression in SARS-CoV-2 infection in vivo. While we hypothesize that elevated serum 90K concentrations in COVID-19 originate from monocytes, our data do not exclude other 90K-producing cell types as additional and/or alternative sources. Strikingly, in scRNA-seq data of respiratory samples of COVID-19 patients, *LGALS3BP* mRNA expression was not upregulated, illustrating the multifaceted ability of SARS-CoV-2 to antagonize or evade cellular sensing machineries in productively infected cells [33,34].

The role of type I IFN in the context of COVID-19 has been discussed controversially and may be time-dependent. On the one hand, overdriven IFN responses are thought to cause severe COVID-19 [35,36]. Since *LGALS3BP* is an ISG, enhanced 90K serum levels in patients with severe COVID-19 could possibly reflect exaggerated type I IFN responses. On the other hand, mounting of an efficient IFN-mediated antiviral state is associated with effective clearance of SARS-CoV-2 infection and milder course of the disease [37]. According to the latter, patients with critical COVID-19 (WHO 6, 7) within the severe disease group (WHO 5-7) had a trend towards lower 90K serum levels than patients classified WHO 5. Similarly, some monocyte fractions of severely ill COVID-19 patients failed to show enhanced *LGALS3BP* upregulation in our scRNA-seq analysis (cohort A), whereas mildly affected patients displayed distinct upregulation. However, this pattern could not be reproduced in cohort B, possibly due to technical differences in the scRNA-seq pipelines and/or unknown clinical differences such as medication or comorbidities. Nevertheless, since *LGALS3BP* is a prototypic ISG [10,38], its upregulation might be merely indicative of the overall mounting of an effective antiviral gene expression program to which *LGALS3BP* is part of, and does not necessarily imply that 90K is an active antiviral component of SARS-CoV-2 infection.

In line with this idea, we failed to establish an anti-SARS-CoV-2 activity of 90K that is exerted directly on SARS-CoV-2 infection. Our study challenges former results proposing 90K as an inhibitor of SARS-CoV-2 spike-mediated membrane fusion [32]. The latter study reported impaired cell-cell membrane fusion of ACE2-positive cells expressing SARS-CoV-2 spike and 90K as well as reduced susceptibility of 90K-overexpressing cells to transduction with lentiviral particles decorated with SARS-CoV-2 spike [39]. However, our cell culture experiments with full-length SARS-CoV-2 failed to identify an inhibitory role of 90K, suggesting that authentic expression, biosynthesis and exposure of SARS-CoV-2 spike on virions is insensitive to restriction by this protein.

In summary, our study describes the expression pattern of *LGALS3BP* in COVID-19 at multiple levels. We propose that 90K/*LGALS3BP* contributes to the global type I IFN response during SARS-CoV-2 infection in vivo without exerting direct antiviral effects detectable in cell culture. Therefore, further investigations on 90K as a potential biomarker and/or potential immunomodulator of SARS-CoV-2 pathogenesis are warranted.

## Data Availability

All data produced in the present study are available upon reasonable request to the authors.
Codes for statistical analyses using linear mixed effects models will be available at https://github.com/GoffinetLab/SARS-CoV-2_90K_patient_study.

## Acknowledgments

B.A. and F.P. are supported by the Charité PhD programme. We thank all participants who took part in NAKO and the staff in this research program.

## Author Contributions

LBJ, BA, CG designed research.

LBJ, BA, JL, GH, JJ, MS, RG performed research.

LBJ, BA, JL, DP, AA, FP, RLC, LK, BM, TJ analyzed data.

AA, LK, TC, DN, JF, TK, TP, JJ, CC, SI, CD, VC, MR, RE, FK, LS, CG supervised research, reviewed and commented on the manuscript.

LBJ, BA, CG wrote the paper.

