## Supplemental Figure 1 for "90K/*LGALS3BP* Expression is Upregulated in COVID-19 but Does Not Restrict SARS-CoV-2 Infection"

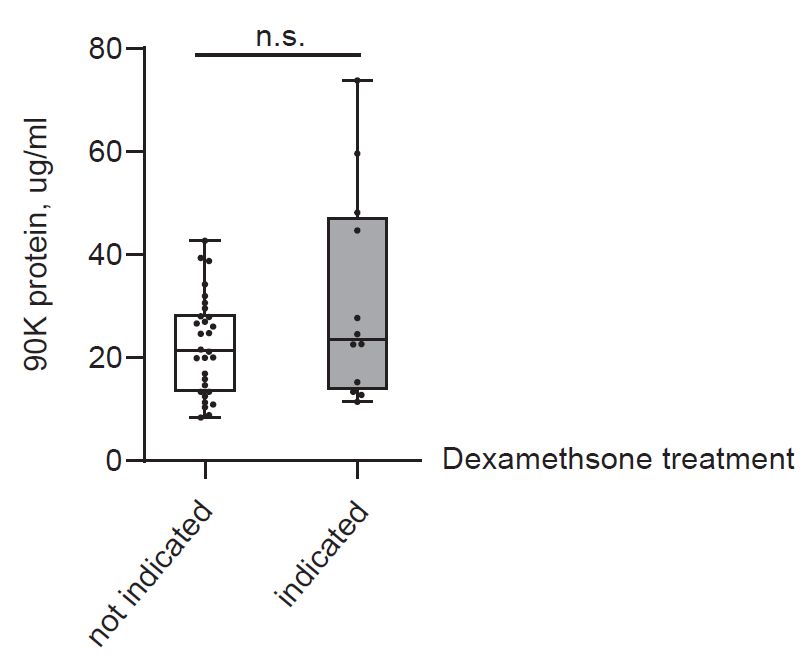

**Suppl. Figure 1. 90K Serum Concentrations Do Not Depend on Dexamethasone Treatment in our COVID-19 Cohort**

90K protein concentrations in serum according to assumed dexamethasone treatment. Dexamethasone was introduced as standard of care for COVID-19 patients with need for oxygen supply in July 2020 (Dexamethasone in Hospitalized Patient...). Patients assigned to the white bar (n=30) were treated for COVID-19 before introduction of dexamethasone as standard of care or, if treated later, presented no need for oxygen supply (WHO 3) and therefore had no indication for dexamethasone treatment. Patients assigned to the gray bar (n=12) were treated from July 2020 on and received oxygen supply (WHO 4 - 7) with indication for dexamethasone treatment. One mean value per patient is depicted. The effect of assumed dexamethasone treatment status on 90K serum levels was assessed using a linear mixed effects model (p=0.07).
