## Supplemental Figure 2 for "90K/*LGALS3BP* Expression is Upregulated in COVID-19 but Does Not Restrict SARS-CoV-2 Infection"

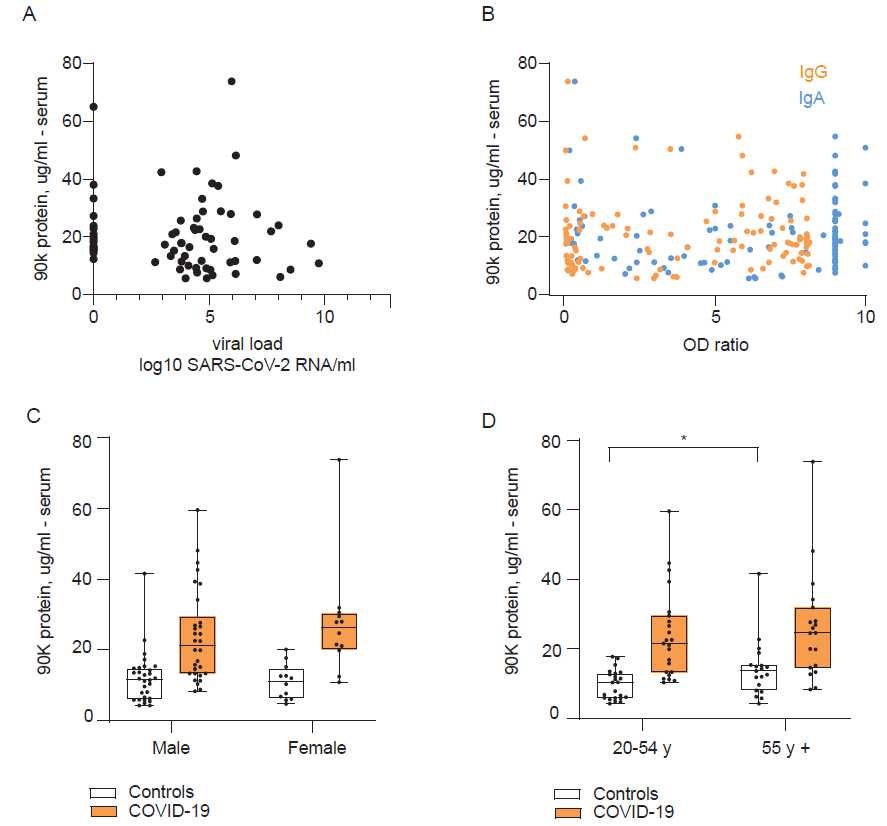

**Suppl. Figure 2. 90K Serum Concentrations in Relation to Infection Status, Antibody Response to SARS-CoV-2, and Demographic Characteristics**

(A) 90K serum protein concentrations and respective SARS-CoV-2 RNA concentrations from nasopharyngeal swabs in COVID-19 patients, n=32 individuals, 68 observations at different time points.

(B) 90K serum protein concentrations and respective IgG/IgA anti-SARS-CoV-2 Spike antibody levels in COVID-19 patients, n=41 individuals, 117 observations at different time points.

(C) 90K protein concentrations in serum and sex of COVID-19 patients and age-matched healthy controls. Male n=30, female n=12, 1 mean value per patient is depicted. The effect of sex on 90K serum concentrations was assessed separately for COVID-19 patients and healthy controls using a linear mixed effects model. This was non-significant in both groups (p=0.39 in COVID-19, p=0.65 in healthy controls).

(D) 90K protein concentrations in serum and age of COVID-19 patients and sex-matched healthy controls. 20-55y n=23, > 55y n=19, 1 mean value per patient is depicted. The effect of age on 90K serum concentrations was assessed separately for COVID-19 patients and healthy controls using a linear mixed effects model. This was non-significant in COVID-19 (p=0.50). For healthy controls this was significant (p=0.027).
