## Supplemental Figure 3 for "90K/*LGALS3BP* Expression is Upregulated in COVID-19 but Does Not Restrict SARS-CoV-2 Infection"

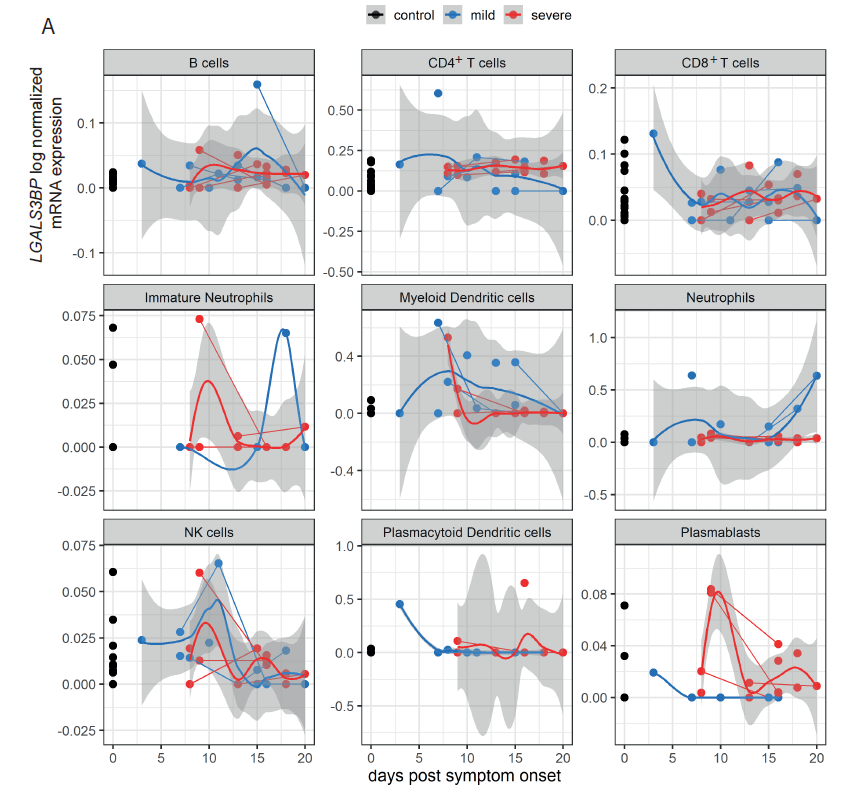

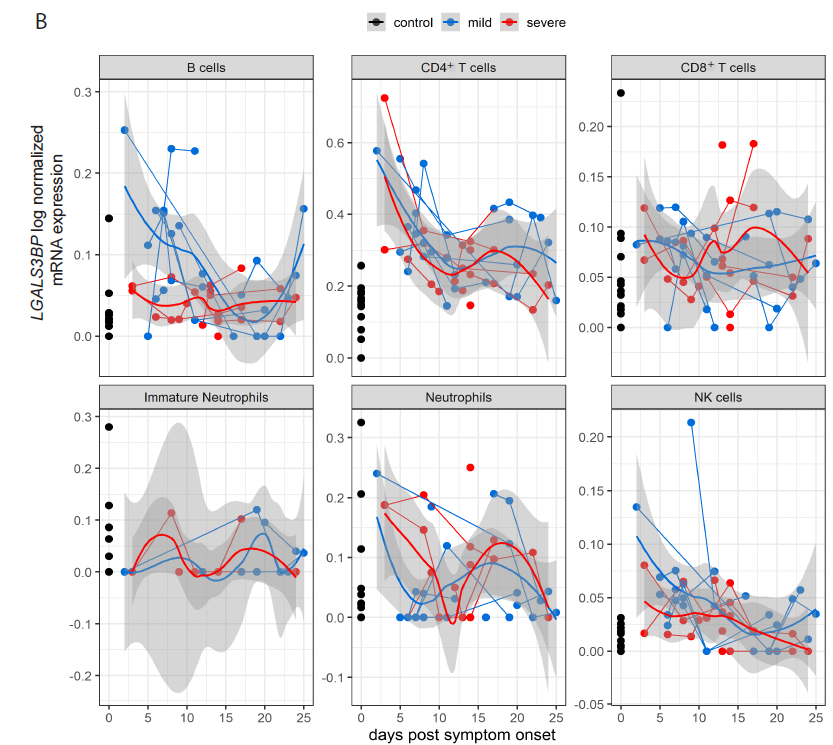

**Suppl. Figure 3. *LGALS3BP* scRNA-seq Analysis from PBMCs, Further Cell Types**

**A**: Cohort A, **B**: Cohort B.

*LGALS3BP* log-normalized expression, assigned to days after symptom onset for each sample. Both cohorts are divided into three groups: Uninfected controls (black), “mild” COVID-19 (defined as WHO grade 2-4 in corresponding publication) (blue) and “severe” COVID-19 (WHO grade 5-7 in corresponding publication) (red). Thick lines indicate smoothed population trends based on a LOESS estimate, thin lines connect subjects. shaded areas indicate 95% CI for the LOESS estimate.

(A) B cells control n=22/22, mild n=8/13, severe n=10/14, CD4^+^ T cells control n=22/22, mild n=8/13, severe n=10/14 , CD8^+^ T cells control n=22/22, mild n=8/13, severe n=10/14, immature neutrophils control n=9/9, mild n=4/4, severe n=10/14, myeloid dendritic cells (mDCs) control n=22/22, mild n=8/13, severe n=10/14, neutrophils control n=19/19 (individuals/time points), mild n=8/11, severe n=10/14, NK cells control n=22/22, mild n=8/13, severe n=10/14 , plasmacytoid dendritic cells (pDCs) control n=22/22, mild n=7/12, severe n=9/14, plasmablasts control n=20/20, mild n=5/6, severe n=10/14

(B) B cells control n=13/13, mild n=8/22, severe n=9/28, CD4^+^ T cells control n=13/13, mild n=8/22, severe n=9/28, CD8^+^ T cells control n=13/13, mild n=8/22, severe n=9/28, immature neutrophils control n=12/12, mild n=5/13, severe n=4/10, neutrophils control n=13/13, mild n=8/21, severe n=9/27, NK cells control n=13/13, mild n=8/22, severe n=9/28.
