## Supplemental Figure 4 for "90K/*LGALS3BP* Expression is Upregulated in COVID-19 but Does Not Restrict SARS-CoV-2 Infection"

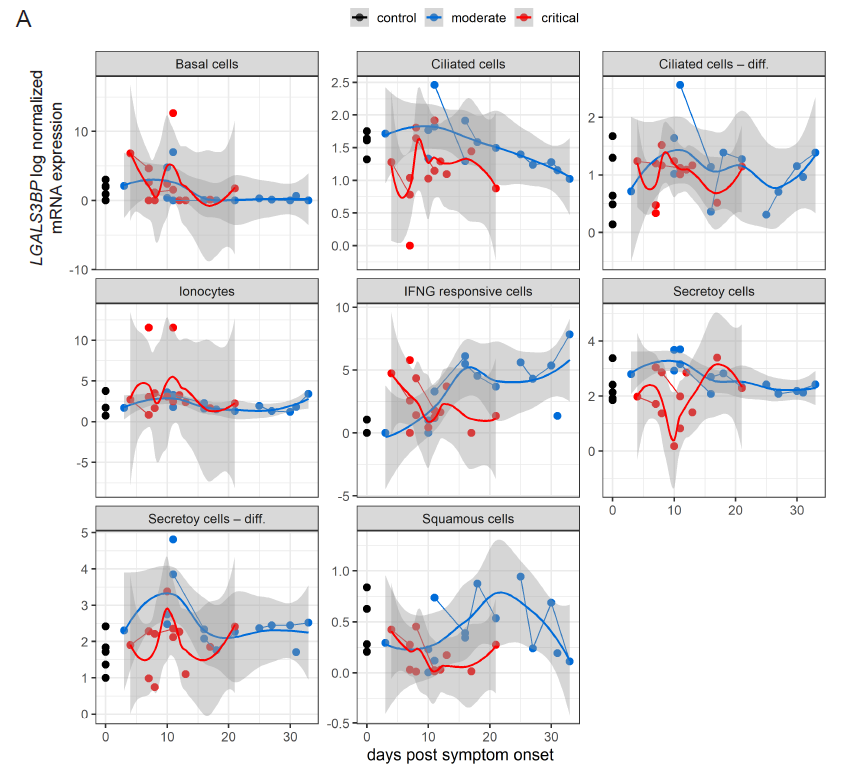

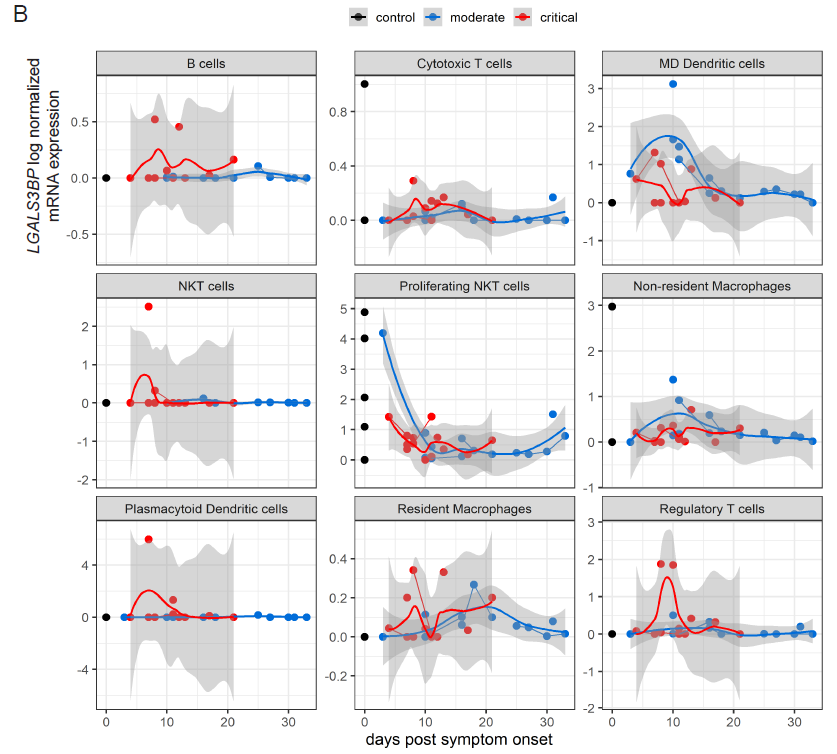

**Suppl. Figure 4. *LGALS3BP* scRNA-seq Analysis from Respiratory Samples**

**A**: Epithelial cells **B**: Immune cells

*LGALS3B*P log-normalized expression at days after symptom onset for each sample. Both cohorts are divided into three groups: Uninfected controls (black), “moderate” COVID-19 (defined as WHO grade 3 in corresponding publication (blue) and “critical” COVID-19 (WHO grade 6-7 in corresponding publication) (red). Repartition of data points for each cell type if not stated otherwise: control n=5/5 (individuals/time points), moderate n=8/14, critical n=11/13. Thick lines indicate smoothed population trends based on a LOESS estimate, thin lines connect subjects. shaded areas indicate 95% CI for the LOESS estimate.

(A) Ionocytes control n=3/3, Secretory cells critical n=10/12, Secretory cells – differentiating critical n=10/12, Squamous cells critical n=9/11.

(B) B cells control n=3/3, moderate n=7/13, critical n=10/11, cytotoxic T cells (CTL) control n=4/4, MD dendritic cells control n=1/1, NKT cells control n=2/2, moderate n=7/13, Plasmacytoid dendritic cells control n=1/1, moderate n=7/13, critical n=8/10, Regulatory T cells control n=4/4, critical n=10/12, critical n=10/12.
