## Supplemental Figure 5 for "90K/*LGALS3BP* Expression is Upregulated in COVID-19 but Does Not Restrict SARS-CoV-2 Infection"

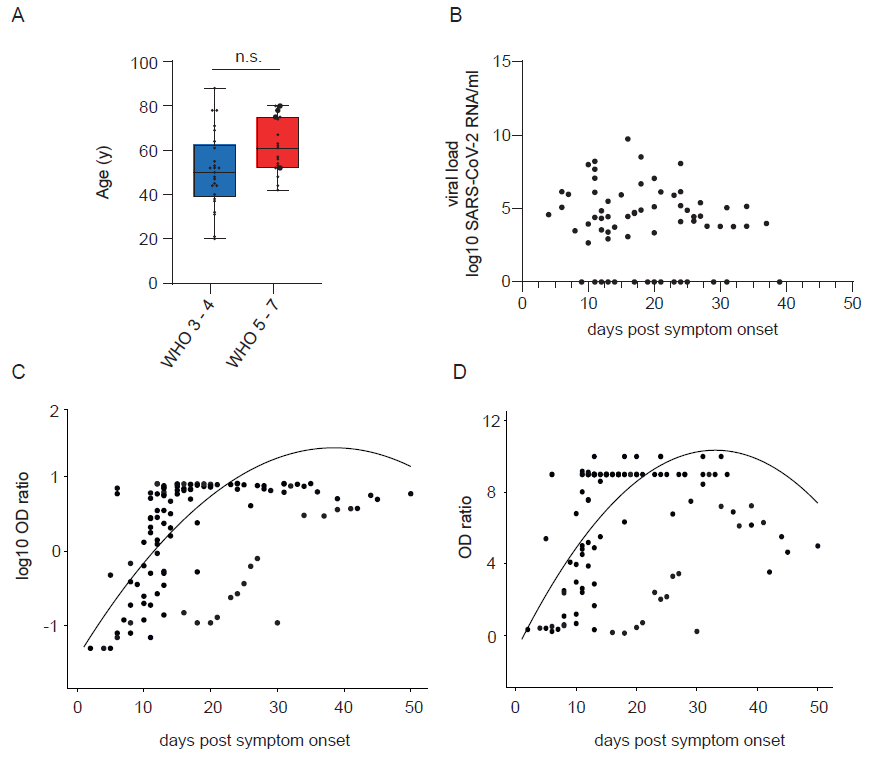

**Suppl. Figure 5. Characterization of COVID-19 Patients**

(A) COVID-19 disease severity and age of infected individuals. WHO 3-4 n=25, WHO 5-7 n=19. Large points indicate deceased patients. Unpaired t-test (p=0.15).

(B) SARS-CoV-2 RNA concentrations in COVID-19 cohort after symptom onset. n=32, 68 time points.

(C) SARS-CoV-2 IgG antibody levels in COVID-19 cohort after symptom onset. n=40, 113 samples at different time points. Log-transformed OD ratios were modelled using a linear mixed effects model, with days post symptom onset (with both a linear and a quadratic term) as fixed effect and patient ID as random intercept. The resulting regression line is indicated (R^2^ = 0.86).

(D) SARS-CoV-2 IgA antibody levels in COVID-19 cohort after symptom onset. n=40, 113 samples at different time points. OD ratios were modelled using a linear mixed effects model, with days post symptom onset (with both a linear and a quadratic term) as fixed effect and patient ID as random intercept. The resulting regression line is indicated (R^2^ = 0.78).
