## Supplemental Figure 6 for "90K/*LGALS3BP* Expression is Upregulated in COVID-19 but Does Not Restrict SARS-CoV-2 Infection"

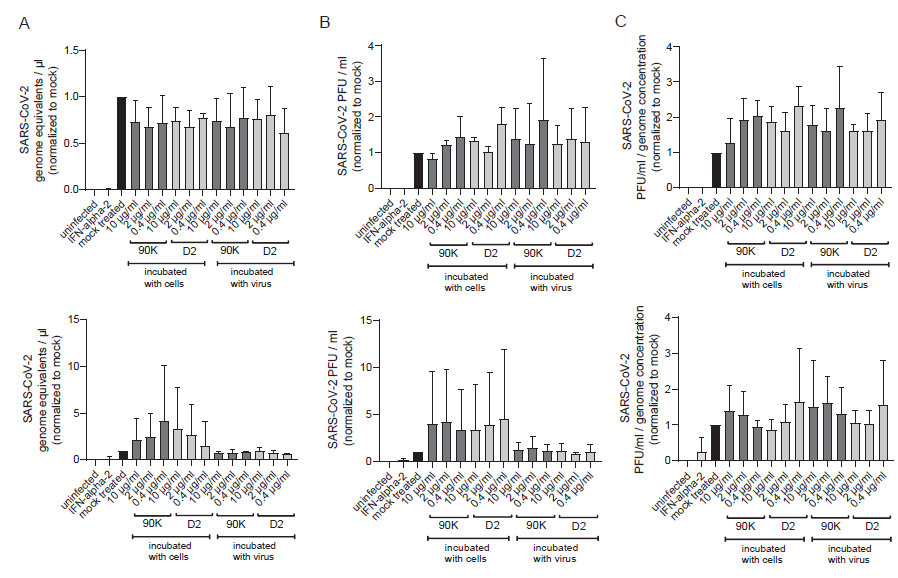

**Suppl. Figure 6. SARS-CoV-2 Particle Release and Infectivity Remains Intact Upon Addition of Exogenous 90K or 90K-D2**

Calu-3 (upper graphs) and Caco-2 (lower graphs) cells or virus stocks were treated with indicated concentrations of purified, full-length 90K and 90K(D2) or left untreated at 37°C for two hours prior to infection. 24 hours post-infection, supernatant was harvested for quantification of SARS-CoV-2 genome equivalents/ul (A) and plaque assays PFU/ml (B) in Vero E6 cells. Data are shown normalized to mock. Particle infectivity was calculated by normalizing SARS-CoV-2 PFU to the genome equivalents/µl, also depicted normalized to mock (C). Results arise from three independent experiments.
