## Supplemental Figure 7 for "90K/*LGALS3BP* Expression is Upregulated in COVID-19 but Does Not Restrict SARS-CoV-2 Infection"

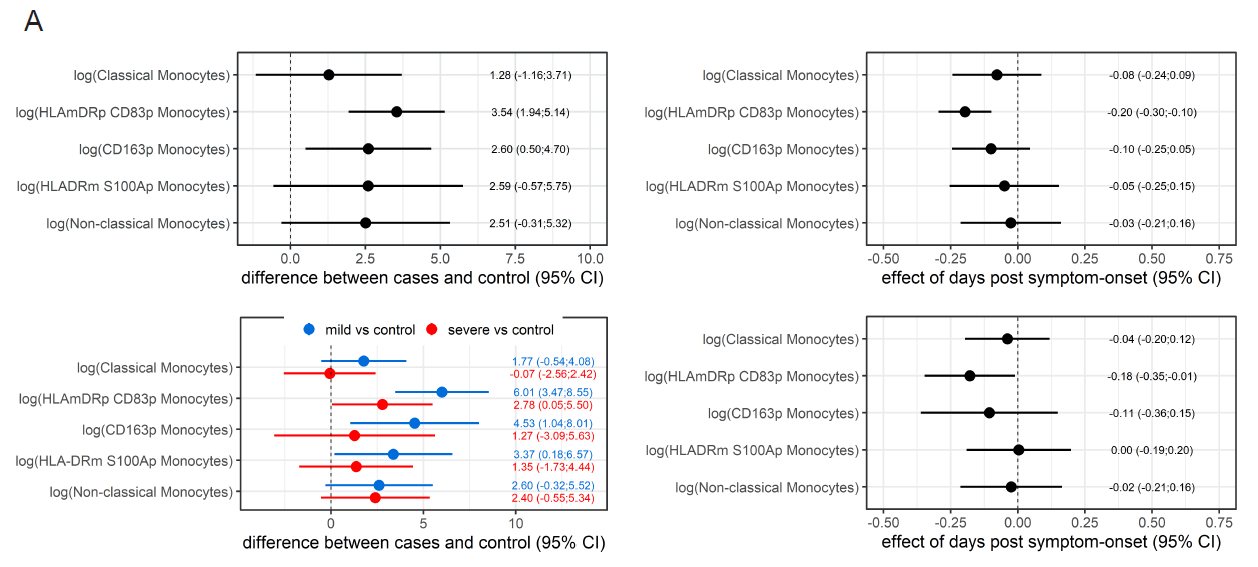

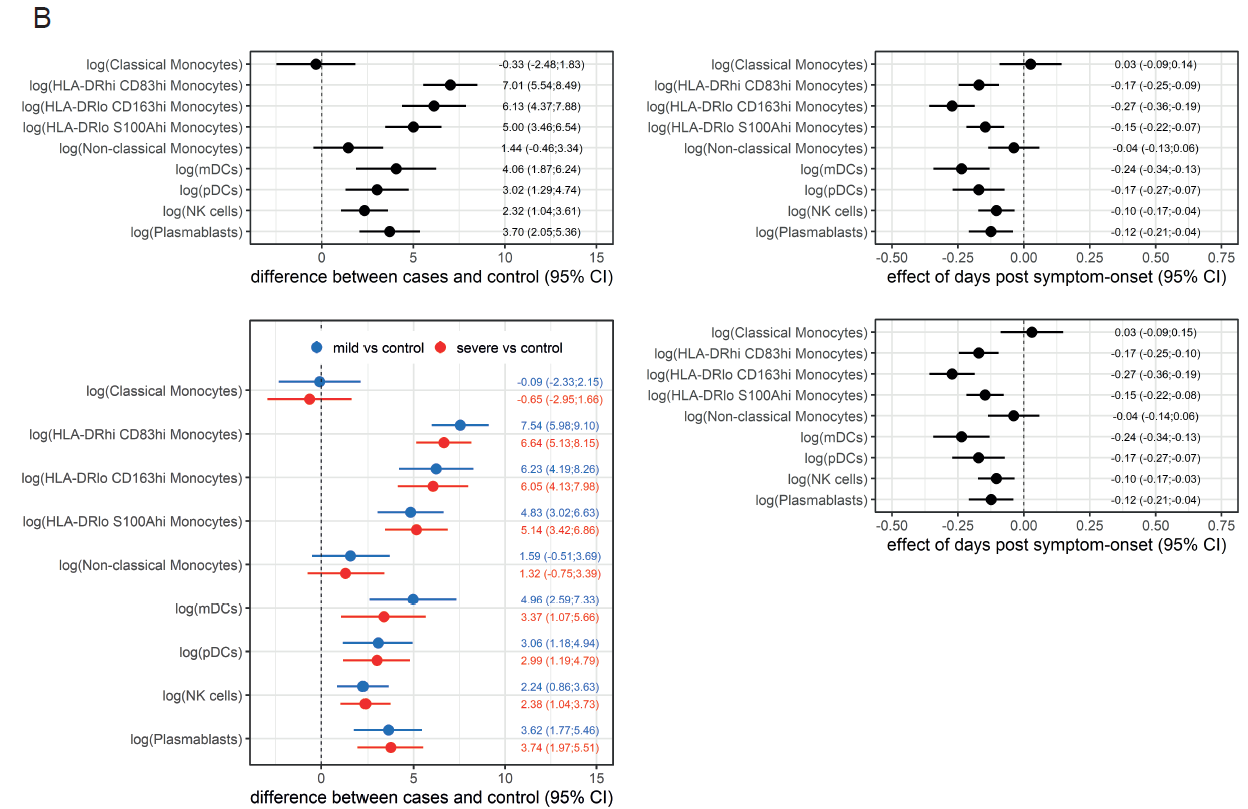

**Suppl. Figure 7. Statistical Analyses of scRNA-seq PBMC Datasets**

Forest plots of analyzed scRNA-seq PBMC datasets from cohort A (A) and cohort B (B). Whiskers indicate 95% CI, derived from mixed linear regression models for two (cases vs. controls, impact of days post symptom onset) and three groups (mild cases vs. controls, severe cases vs. controls, impact of days post symptom onset).
