## Supplemental Table 1 for "90K/*LGALS3BP* Expression is Upregulated in COVID-19 but Does Not Restrict SARS-CoV-2 Infection"

**Suppl. Table 1. Overview of Sampled Data in COVID-19 Cohort**

Each patient sample is attributed to sample type, disease severity, sex (M = male, F = female), age in ten-year sections and days post symptom onset on sampling day.

| Patient N° | Sample | 90K Serum ELISA | 90K PBMC ELISA | *LGALS3BP* qRT PCR | WHO Grade | Sex | Age section (y) | Days post Symptom Onset |
| --- | --- | --- | --- | --- | --- | --- | --- | --- |
| **1** | **A** | **X** |  |  | **3** | **M** | **<25** | **16** |
| **1** | **B** | **X** |  |  | **3** | **M** | **<25** | **18** |
| **1** | **C** | **X** |  |  | **3** | **M** | **<25** | **20** |
| **1** | **D** | **X** |  |  | **3** | **M** | **<25** | **21** |
| **1** | **E** | **X** |  |  | **3** | **M** | **<25** | **23** |
| **1** | **F** | **X** |  |  | **3** | **M** | **<25** | **24** |
| **1** | **G** | **X** |  |  | **3** | **M** | **<25** | **25** |
| **1** | **H** | **X** |  |  | **3** | **M** | **<25** | **26** |
| **1** | **I** | **X** |  |  | **3** | **M** | **<25** | **27** |
| **1** | **J** | **X** |  |  | **3** | **M** | **<25** | **30** |
| **1** | **K** | **X** |  |  | **3** | **M** | **<25** | **32** |
| **1** | **L** | **X** |  |  | **3** | **M** | **<25** | **34** |
| **1** | **M** | **X** |  |  | **3** | **M** | **<25** | **37** |
| **1** | **N** | **X** |  |  | **3** | **M** | **<25** | **39** |
| **1** | **O** | **X** |  |  | **3** | **M** | **<25** | **41** |
| **2** | **A** | **X** |  |  | **3** | **F** | **25-34** | **8** |
| **2** | **B** | **X** |  |  | **3** | **F** | **25-34** | **10** |
| **2** | **C** | **X** |  |  | **3** | **F** | **25-34** | **11** |
| **2** | **D** | **X** |  |  | **3** | **F** | **25-34** | **12** |
| **2** | **E** | **X** |  |  | **3** | **F** | **25-34** | **13** |
| **3** | **A** | **X** |  |  | **3** | **M** | **45-54** | **11** |
| **3** | **B** | **X** |  |  | **3** | **M** | **45-54** | **12** |
| **3** | **C** | **X** |  |  | **3** | **M** | **45-54** | **13** |
| **3** | **D** | **X** |  |  | **3** | **M** | **45-54** | **16** |
| **3** | **E** | **X** |  |  | **3** | **M** | **45-54** | **18** |
| **3** | **F** | **X** |  |  | **3** | **M** | **45-54** | **20** |
| **3** | **G** | **X** |  |  | **3** | **M** | **45-54** | **23** |
| **3** | **H** | **X** |  |  | **3** | **M** | **45-54** | **25** |
| **4** | **A** | **X** |  |  | **4** | **M** | **65+** | **13** |
| **4** | **B** | **X** |  |  | **4** | **M** | **65+** | **14** |
| **4** | **C** | **X** |  |  | **4** | **M** | **65+** | **17** |
| **4** | **D** | **X** |  |  | **4** | **M** | **65+** | **19** |
| **5** | **A** | **X** |  |  | **4** | **M** | **25-34** | **8** |
| **5** | **B** | **X** |  |  | **4** | **M** | **25-34** | **11** |
| **5** | **C** | **X** |  |  | **4** | **M** | **25-34** | **12** |
| **5** | **D** | **X** |  |  | **4** | **M** | **25-34** | **15** |
| **5** | **E** | **X** |  |  | **4** | **M** | **25-34** | **17** |
| **6** | **A** | **X** |  |  | **7** | **M** | **55-64** | **24** |
| **6** | **B** | **X** |  |  | **7** | **M** | **55-64** | **26** |
| **6** | **C** | **X** |  |  | **7** | **M** | **55-64** | **28** |
| **6** | **D** | **X** |  |  | **7** | **M** | **55-64** | **31** |
| **6** | **E** | **X** |  |  | **7** | **M** | **55-64** | **33** |
| **6** | **F** | **X** |  |  | **7** | **M** | **55-64** | **35** |
| **7** | **A** | **X** |  |  | **6** | **F** | **65+** | **6** |
| **7** | **B** | **X** |  |  | **6** | **F** | **65+** | **8** |
| **7** | **C** | **X** |  |  | **6** | **F** | **65+** | **11** |
| **7** | **D** | **X** |  |  | **6** | **F** | **65+** | **13** |
| **7** | **E** | **X** |  |  | **6** | **F** | **65+** | **15** |
| **8** | **A** | **X** |  |  | **3** | **F** | **45-54** | **10** |
| **8** | **B** | **X** |  |  | **3** | **F** | **45-54** | **11** |
| **9** | **A** | **X** |  |  | **5** | **M** | **55-64** | **12** |
| **9** | **B** | **X** |  |  | **5** | **M** | **55-64** | **17** |
| **9** | **C** | **X** |  |  | **5** | **M** | **55-64** | **19** |
| **10** | **A** | **X** |  |  | **6** | **M** | **65+** | **12** |
| **10** | **B** | **X** |  |  | **6** | **M** | **65+** | **13** |
| **10** | **C** | **X** |  |  | **6** | **M** | **65+** | **16** |
| **10** | **D** | **X** |  |  | **6** | **M** | **65+** | **18** |
| **10** | **E** | **X** |  |  | **6** | **M** | **65+** | **20** |
| **11** | **A** | **X** |  |  | **7** | **M** | **45-54** | **8** |
| **12** | **A** | **X** |  |  | **6** | **M** | **65+** | **4** |
| **12** | **B** | **X** |  |  | **6** | **M** | **65+** | **6** |
| **12** | **C** | **X** |  |  | **6** | **M** | **65+** | **8** |
| **13** | **A** | **X** |  |  | **3** | **F** | **55-64** | **21** |
| **14** | **A** | **X** |  |  | **3** | **M** | **45-54** | **9** |
| **14** | **B** | **X** |  |  | **3** | **M** | **45-54** | **11** |
| **14** | **C** | **X** |  |  | **3** | **M** | **45-54** | **13** |
| **15** | **A** | **X** |  |  | **6** | **F** | **45-54** | **11** |
| **15** | **B** | **X** |  |  | **6** | **F** | **45-54** | **13** |
| **15** | **C** | **X** |  |  | **6** | **F** | **45-54** | **15** |
| **16** | **A** | **X** |  |  | **6** | **M** | **45-54** | **11** |
| **16** | **B** | **X** |  |  | **6** | **M** | **45-54** | **13** |
| **16** | **C** | **X** |  |  | **6** | **M** | **45-54** | **15** |
| **17** | **A** | **X** |  |  | **5** | **M** | **55-64** | **14** |
| **17** | **B** | **X** |  |  | **5** | **M** | **55-64** | **16** |
| **17** | **C** | **X** |  |  | **5** | **M** | **55-64** | **18** |
| **18** | **A** | **X** |  |  | **3** | **F** | **35-44** | **asymptomatic** |
| **18** | **B** | **X** |  |  | **3** | **F** | **35-44** | **asymptomatic** |
| **18** | **C** | **X** |  |  | **3** | **F** | **35-44** | **asymptomatic** |
| **19** | **A** | **X** |  |  | **4** | **M** | **55-64** | **12** |
| **19** | **B** | **X** |  |  | **4** | **M** | **55-64** | **14** |
| **20** | **A** | **X** |  |  | **5** | **M** | **55-64** | **10** |
| **20** | **B** | **X** |  |  | **5** | **M** | **55-64** | **12** |
| **21** | **A** | **X** |  |  | **3** | **F** | **35-44** | **13** |
| **22** | **A** | **X** |  |  | **4** | **M** | **45-54** | **12** |
| **22** | **B** | **X** |  |  | **4** | **M** | **45-54** | **14** |
| **23** | **A** | **X** |  |  | **3** | **M** | **45-54** | **5** |
| **24** | **A** | **X** |  |  | **3** | **F** | **35-44** | **12** |
| **25** | **A** | **X** |  |  | **3** | **M** | **65+** | **13** |
| **26** | **A** | **X** | **X** | **X** | **4** | **M** | **65+** | **20** |
| **27** | **A** | **X** | **X** | **X** | **3** | **F** | **65+** | **6** |
| **28** | **A** | **X** |  |  | **4** | **M** | **55-64** | **6** |
| **29** | **A** | **X** |  | **X** | **3** | **F** | **<25** | **15** |
| **30** | **A** | **X** |  |  | **3** | **F** | **35-44** | **2** |
| **31** | **A** | **X** | **X** | **X** | **7** | **M** | **65+** | **29** |
| **31** | **B** | **X** |  |  | **7** | **M** | **65+** | **36** |
| **31** | **C** | **X** |  |  | **7** | **M** | **65+** | **44** |
| **31** | **D** | **X** |  |  | **7** | **M** | **65+** | **50** |
| **32** | **A** | **X** | **X** | **X** | **7** | **M** | **65+** | **24** |
| **32** | **B** | **X** | **X** |  | **7** | **M** | **65+** | **31** |
| **32** | **C** | **X** |  | **X** | **7** | **M** | **65+** | **39** |
| **32** | **D** | **X** |  |  | **7** | **M** | **65+** | **45** |
| **32** | **E** | **X** | **X** | **X** | **7** | **M** | **65+** | **42** |
| **33** | **A** | **X** |  |  | **5** | **M** | **45-54** | **5** |
| **34** | **A** | **X** | **X** | **X** | **4** | **W** | **65+** | **7** |
| **35** | **A** | **X** | **X** | **X** | **3** | **M** | **45-54** | **13** |
| **36** | **A** | **X** | **X** | **X** | **6** | **M** | **35-44** | **17** |
| **36** | **B** | **X** | **X** |  | **6** | **M** | **35-44** | **24** |
| **36** | **C** | **X** | **X** |  | **6** | **M** | **35-44** | **31** |
| **37** | **A** | **X** |  |  | **7** | **M** | **55-64** | **10** |
| **37** | **B** | **X** | **X** | **X** | **7** | **M** | **55-64** | **17** |
| **37** | **C** | **X** |  | **X** | **7** | **M** | **55-64** | **24** |
| **38** | **A** | **X** |  | **X** | **4** | **M** | **45-54** | **11** |
| **38** | **B** | **X** | **X** | **X** | **4** | **M** | **45-54** | **18** |
| **39** | **A** | **X** |  |  | **4** | **M** | **35-44** | **28** |
| **40** | **A** | **X** | **X** | **X** | **3** | **M** | **45-54** | **10** |
| **41** | **A** | **X** |  | **X** | **7** | **M** | **45-54** | **13** |
| **41** | **B** | **X** |  |  | **7** | **M** | **45-54** | **20** |
| **42** | **A** | **X** | **X** | **X** | **7** | **M** | **65+** | **27** |
| **42** | **B** | **X** | **X** | **X** | **7** | **M** | **65+** | **34** |
| **43** | **A** |  |  | **X** | **5** | **W** | **35-44** | **16** |
| **44** | **A** |  | **X** |  | **5** | **W** | **65+** | **10** |
