## Supplemental Methods Text for "90K/*LGALS3BP* Expression is Upregulated in COVID-19 but Does Not Restrict SARS-CoV-2 Infection"

**Classification of Disease Severity**

We assessed COVID-19 disease severity using the World Health Organization (WHO) ordinal scale for clinical improvement on sampling day [[40]](https://paperpile.com/c/oA8C2P/mEO9).

WHO 1: Asymptomatic infection

WHO 2: Symptomatic infection, ambulatory care

WHO 3: Hospitalization, no supplemental oxygen required

WHO 4: Supplemental oxygen required

WHO 5: High Flow or Continuous Positive Airway Pressure (CPAP) required

WHO 6: Mechanical ventilation required

WHO 7: Extracorporeal Membrane Oxygenation (ECMO) and/or Continuous Renal

Replacement Therapy (CRRT) required

WHO 8: Death

For practical reasons, we designated patients admitted to general hospital wards as “mild COVID-19” (WHO 3-4) and patients admitted to intensive care units as “severe COVID-19” (WHO 5-7). Within the latter, patients receiving mechanical ventilation, ECMO or CRRT were designated as “critical COVID-19” (WHO 6-7). Patients with multiple WHO grades within the sampling period (6 out of 42 patients) were assigned the highest disease severity grade.

**Healthy Controls**

A control population was provided by the study center Berlin-Mitte of the German National Cohort (Gesundheitsstudie, NAKO*) –* Germany’s largest population-based longitudinal cohort study (Kühn et al. 2014; German National Cohort (GNC) Consorti...; Schipf et al. 2020). 42 serum samples from uninfected individuals were selected randomly from participants recruited in the study region (Berlin), after exclusion of potentially confounding conditions that may lead to an elevation of 90K serum levels, as specified in the study patients with COVID-19. To rule out SARS-CoV-2 infection in the controls, we chose serum samples collected between 2014 and 2015. Samples were requested according to Use&Access regulations of the NAKO. Control samples were manually matched 1:1 to our COVID-19 patients for sex and age categories: <25 years, 25 - 34 years, 35 - 44 years, 45 - 54 years, 55 - 64 years, ≥65 years. Details on assignment of healthy controls to COVID-19 patients are provided in Suppl. Table 2. Retrieval of referred blood samples was approved by the ethics committee of Charité - Universitätsmedizin Berlin (EA1/076/13 vom 28.3.2014).

Retrieval of further blood samples and cell isolation from healthy anonymized donors was conducted with approval of the local ethics committee (Ethical review committee of Charité Berlin, vote 3674-2017).

**Analysis of Single Cell RNA-sequencing Data of PBMCs**

Analyzed single cell RNA-sequencing (scRNA-seq) data from PBMCs originate from a published Dual Center Cohort Study [[27]](https://paperpile.com/c/oA8C2P/Tldj). Two patient cohorts had previously been recruited between March and July 2020. PBMCs from Cohort A (27 samples from 18 SARS-CoV-2 patients - mild (WHO 2-4): n=8, severe (WHO 5-7): n=10, and 22 healthy controls) had been collected at Charité-Universitätsmedizin Berlin. Data had been generated by scRNA-seq with a droplet-based platform. PBMCs from Cohort B (50 samples from 17 SARS-CoV-2 patients - mild: n=8, severe: n=9, 13 healthy controls) had been collected at University Hospital Bonn and scRNA-seq data had been generated by a microwell-based system. As the scRNA-seq data of both cohorts had been generated by different experimental approaches, we analyzed each cohort separately in this study. Pre-processed scRNA-seq datasets were obtained from the European Genome Archive (EGA) under access number EGAS00001004571 and data were extracted using the FetchData function of the R package Seurat [[41]](https://paperpile.com/c/oA8C2P/iHbv).

**Analysis of Single Cell RNA-sequencing Data of Respiratory Samples**

We analyzed a published dataset of 32 respiratory samples from 19 hospitalized COVID-19 patients and five healthy controls for *LGALS3BP* expression, originating from a published scRNA-seq study [[28]](https://paperpile.com/c/oA8C2P/YVxL). Analyzed samples had been obtained through nasopharyngeal swabs, bronchial protected specimen brushes, and bronchoalveolar lavages and had been processed with a droplet-based system. Amongst the 19 COVID-19 patients, eight had a moderate disease course (WHO 3), while eleven had a critical disease course (WHO 6-7). The pre-processed scRNA-seq dataset was obtained from the European Genome Archive (EGA) under access number EGAS00001004481 and data were extracted using the *FetchData* function of the R package Seurat [[41]](https://paperpile.com/c/oA8C2P/iHbv)*.*

**Transfection**

HEK293T/ACE2 cells were transfected with empty vector (pcDNA.6myc) or pcDNA.90K-myc [[10]](https://paperpile.com/c/oA8C2P/1Cfwn) by calcium-phosphate transfection using TaKaRa CalPhos™ Mammalian Transfection Kit following the manufacturer's instructions.

**Reagents**

Roferon-A (IFN-α2a) was purchased from Roche (Basel, Switzerland) and used at a concentration of 500 IU/ml. Remdesivir (Gilead Sciences) was kindly provided by the Department of Infectious Diseases and Respiratory Medicine, Charité - Universitätsmedizin Berlin and used at a concentration of 10 µM.

**Purification of 90K and 90K-D2**

Individual cDNAs encoding full 90K and e 90K-D2 were generated by polymerase chain reaction (PCR). D2 is defined as a 28 kDa fragment corresponding to amino acid residues 134-288 of 90K. Fragments were subcloned into an Evitria’s proprietary vector system (EVITRIA, Schlieren, Switzerland) allowing fusion in-frame to the 6xHis tag sequence C-terminally to the domain.

90K and 90K-D2 were transiently expressed by transfection into Chinese Hamster Ovary (CHO) cells. Concentrated culture supernatants were applied to a column of Ni-NTA Superflow (Qiagen) and proteins were eluted with increasing concentrations of imidazole according to the manufacturer’s instructions.

**SARS-CoV-2 Plaque Titration Assay**

The infectious titer was calculated via plaque titration assay. Vero E6 cells were plated at 3.5 x 10^5^ cell/ml in 24 wells and infected with 200 µl of a serial dilution of virus-containing cell culture supernatant diluted in OptiPro serum-free medium. One hour after adsorption, supernatants were removed and cells overlaid with 2.4% Avicel (FMC BioPolymers) mixed 1:1 in 2 x DMEM. Three days post-infection, the overlay was removed, cells were fixed in 6% formaldehyde and stained with a 0.2% crystal violet, 2% ethanol and 10% formaldehyde.

**Anti-90K ELISA**

*Sera.* Sera were analyzed with the Thermo Fisher Scientific s90K/Mac-2BP ELISA Kit according to the manufacturer’s instructions. Sera were analyzed in duplicates in 1:100 and 1:1000 dilutions.

*PBMCs.* Frozen PBMC pellets (1,5 - 9 x 10^6^ cells) were lysed in 1% Triton X-100 and applied to Thermo Fisher Scientific s90K/Mac-2BP ELISA Kit following the manufacturer’s instructions in a 1:14 dilution. PBMCs were analyzed in single measurements as the amount of patient material was not sufficient for analysis in duplicates.

**Anti-SARS-CoV-2 IgA/IgG ELISA**

Human sera were examined for presence of specific antibodies (IgG and IgA) to the S1 subdomain of The SARS-CoV-2 spike protein using an ELISA kit (Euroimmun, Lübeck, Germany) as described before [[42,43]](https://paperpile.com/c/oA8C2P/kNDMb+7xG8). Samples were tested at a 1:101 dilution and results were considered positive above an optical density (OD) ratio of 1.1. Automated measurement was performed with Euroimmun Analyzer I.

**Viral RNA Load Analysis from Nasopharyngeal Swabs**

SARS-CoV-2 RNA quantification was performed by real-time RT-PCR from upper respiratory tract swabs obtained within standard care. RNA concentrations were quantified by RT-qPCR targeting SARS-CoV-2 E gene [[44]](https://paperpile.com/c/oA8C2P/AbpWG) and are given as logarithm base 10 of the number of RNA copies (viral load) per ml using an empirical formula derived from testing standard curves of SARS-CoV-2 RNA and cell culture supernatants [[45]](https://paperpile.com/c/oA8C2P/y3clc).

**qRT-PCR**

Total RNA extraction from PBMCs and cell lines was performed with Qiagen RNeasy Micro Kit according to the manufacturer’s instructions. RNA isolation was followed by 2-step PCR. Quantification was performed by RT-qPCR in Roche LightCycler 480 II using TaqMan PCR technology with a premade primer-probe kit for *LGALS3BP, MX2*, and *IFIT1* (Applied Biosystems). For *LGALS3BP*, oligonucleotide primers 5′-GCTTCCTTCCTCTCTGCAATGA-3′ (forward), 5′-TCAGGTGAGTAGGGCGACATC-3′ (reverse), 5′-FAM-CTTCAACAACCGGCCAC-TAMRA-3′ (fluorescent probe) were used. For *MX2* and *IFIT1*, we used assay IDs Hs01550814_m1 and Hs01911452_s1, respectively (Thermo Fisher Scientific Waltham, Massachusetts, USA).

Relative mRNA levels were determined using the ΔCT method, with human *RNASEP* mRNA (Applied Biosystems) as internal reference. Each sample was analyzed in triplicate. Viral RNA extraction was performed using Macherey Nagel Nucleospin RNA Virus Mini Kit. SARS-CoV-2 genome equivalents were quantified by RT-qPCR targeting SARS-CoV-2 E gene using the following primers: E_Sarbeco_F: ACAGGTACGTTAATAGTTAATAGCGT; E_Sarbeco_P1: FAM-ACACTAGCCATCCTTACTGCGCTTCG-BBQ; E_Sarbeco_R: ATATTGCAGCAGTACGCACACA [[44]](https://paperpile.com/c/oA8C2P/AbpWG). RT-qPCRs were performed using the Superscript III OneStep RT-PCR kit (Invitrogen, Darmstadt, Germany). Absolute quantification was performed using SARS-CoV-2-specific in vitro-transcribed RNA standards. Data analysis was performed using Roche LightCycler 480 1.5.1.62SP3 software.

**Immunoblotting**

Cell lysates were generated with M-PER Mammalian Protein Extraction Reagent (Thermo Fisher Scientific, Waltham, Massachusetts, USA). For virus-containing supernatant, 1x SDS-buffer was added to the sample. Proteins were separated on a 10% SDS-PAGE and transferred onto nitrocellulose using a semi-dry transfer system (Bio-Rad Laboratories, Hercules, California, USA). Membranes were blocked with 5% milk powder solution for one hour and incubated overnight with the following primary antibodies: rabbit anti-myc (71D10, Cell Signaling Technology, Danvers, USA), goat anti-Galectin-3BP/MAC-2BP (AF2226, R&D Systems, Minnesota, USA), goat anti-MX2 (sc-47197, Santa Cruz Biotechnology, California, USA), mouse anti-IFIT1 (CF500948, Origene, Maryland, USA), rabbit anti-nucleocapsid (GTX135361, Biozol, Eching, Germany), mouse anti-ACE2 (10108-MM36, Sino Biological, Beijing, China), rabbit anti-SARS Spike (NB100-56578, Novus Biologicals, Minneapolis, USA)**,** rabbit anti-α-tubulin (#2144, Cell Signaling Technology, Massachusetts, USA), and mouse-anti human actin (#8226, Abcam, Cambridge, UK). Secondary antibodies conjugated to Alexa 680/800 fluorescent dyes were used for detection and quantification by Odyssey Infrared Imaging System (LI-COR Biosciences Lincoln, NE, USA).

**Flow Cytometry**

Cells were fixed in 4% PFA (Carl Roth) and permeabilized with 0.1% Triton X-100 (Thermo Fisher Scientific) in PBS before immunostaining with the following antibodies: rabbit anti-nucleocapsid (GTX135361, Biozol, Eching, Germany), goat anti-Galectin-3BP/MAC-2BP (AF2226, R&D Systems, Minnesota, USA), and mouse anti-ACE2 (10108-MM36, Sino Biological, Beijing, China). Secondary antibodies conjugated to Alexa Fluor 488 or 647 (1:1,500; Invitrogen) were used for detection. Flow cytometry analysis was performed using FACS Celesta with BD Diva Software (BD Biosciences) and FlowJo V10 Software (FlowJo).
